# Barriers and enablers to influenza vaccination uptake in adults with chronic respiratory conditions: Applying the behaviour change wheel to specify multi-levelled tailored intervention content

**DOI:** 10.1101/2020.11.18.20233783

**Authors:** Allyson J Gallant, Paul Flowers, Karen Deakin, Nicola Cogan, Susan Rasmussen, David Young, Lynn Williams

**Affiliations:** University of Strathclyde, Scotland, United Kingdom

**Author notes:** **Corresponding Author.** Contact Details: Dr Lynn Williams, School of Psychological Sciences & Health, University of Strathclyde, 40 George Street, Glasgow, G1 1QE, Scotland, UK.

**Keywords:** influenza vaccination, vaccine hesitancy, behaviour change wheel, theoretical domains framework, behaviour change techniques

## Abstract

**Objectives:** To specify future intervention content to enhance influenza vaccination uptake using the Behaviour Change Wheel (BCW).

**Design:** Cross-sectional, multi-modal data collection and subsequent behaviourally informed analysis and expert stakeholder engagement.

**Methods:** Content analysis was initially used to identify barriers and enablers to influenza vaccination from nine semi-structured focus groups, 21 individual interviews and 101 open-ended survey responses. Subsequently, the Theoretical Domains Framework (TDF) and the BCW were used to specify evidence-based and theoretically-informed future intervention content in the form of preliminary recommendations. Finally, drawing on the APEASE criteria, expert stakeholders refined our recommendations to yield a range of multi-levelled potentially actionable ideas.

**Results:** The TDF domain of ‘Beliefs about Consequences’ was the most frequently mapped domain with themes relating to ‘perceptions of side effects (barrier)’ and ‘feeling protected from catching flu (enabler)’. The next most important domain was ‘Environmental Context and Resources’ with themes relating to ‘time constraints (barrier)’ and ‘receiving reminders to vaccinate (enabler)’. Next, ‘Social Influences’ was identified with themes relating to ‘encouragement from others (enabler)’, followed by ‘Emotion’ with themes relating to ‘fear of needles (barrier)’. These factors mapped to seven of the nine intervention functions and 22 identified behaviour change techniques (BCTs). Stakeholders reduced an initial 26 recommendations to 21.

**Conclusions:** Our comprehensive analyses showed that the factors affecting vaccine uptake were multifaceted and multileveled. The study suggested a suite of complementary multi-level intervention components may usefully be combined to enhance vaccination uptake involving a range of diverse actors, intervention recipients and settings.

**Statement of Contribution:** *What is already known on this subject?:*

- Uptake of the influenza vaccination in those with an “at-risk” health condition is low and has been decreasing year on year.
- The reasons for vaccine hesitancy are complex and involve psychological, social and contextual factors.
- There is a lack of theory-based intervention content aimed at increasing influenza vaccination uptake.

*What does this study add?:*

- This study showed that the factors affecting vaccine uptake were multifaceted and multileveled. They could be theorised as relating to the TDF domains of ‘Beliefs about Consequences’, ‘Environmental Context and Resources’, ‘Social Influences’ and ‘Emotion’.
- With the help of key stakeholders the study suggested a suite of complementary multi-level intervention components may be most useful to enhance vaccination compliance involving a range of diverse actors, intervention recipients and settings.
- Mass and social media interventions, and interactions between recipients and healthcare providers should include clear and concise information about vaccine side-effects and directly address misinformation. Community-based vaccination delivery methods should be enhanced by modifying traditional and adopting novel approaches.

## Introduction

Seasonal influenza remains a significant public health threat. The most effective way of preventing seasonal influenza is through vaccination, which is offered to particular at-risk groups (e.g. individuals with chronic medical conditions). However, a large number of those eligible to receive the seasonal influenza vaccination decide against vaccination. The term ‘vaccine hesitancy’ refers to the ‘delay in acceptance or refusal of vaccines despite availability of vaccine services’ (MacDonald & the SAGE Working Group on Vaccine Hesitancy, 2015). Seasonal influenza vaccination uptake, even among those who are at high risk (e.g. those with chronic illnesses), is typically less than 50% in most European countries, which falls substantially below the World Health Organisation target of 75% (Jorgensen et al., 2018). In Scotland, where the present study was conducted, seasonal influenza vaccine uptake has declined considerably in recent years in the under 65 year-old clinical risk group (i.e. those who are under the age of 65 but receive the vaccination due to an underlying chronic medical condition). Specifically, within the chronic respiratory disease group, who form the largest proportion of this under-65 clinical risk group, uptake was 44.6% in 2018-19 (Health Protection Scotland, 2020). It is therefore necessary for research to examine the barriers and enablers to vaccine acceptance in order to inform strategies to address vaccine hesitancy (Brewer et al., 2017).

The reasons for vaccine hesitancy are complex and involve psychological, social and contextual factors (Brewer et al., 2017). Sociodemographic factors, such as age, education and ethnicity, have been inconsistently linked to uptake (Schmid et al., 2017). Contextual factors, relating to how the vaccination programme is delivered and the resulting practical barriers that individuals face in terms of the convenience of getting vaccinated also play a role. However, there is increasing recognition of the role that psychological processes, such as perceived susceptibility and perceived severity, and social norms, play in shaping vaccination behaviour (Schmid et al., 2017).

In their review, Schmid et al. (2017) identified that psychological theories have rarely been applied in understanding the psychological factors influencing influenza vaccine uptake. In those studies that do use theoretically-derived psychological factors, the vast majority of studies have used the Health Belief Model (Borthwick et al., 2020). Applying theory in the design of multi-component interventions has been shown to improve the intervention effectiveness (Craig et al., 2008). To simplify the use of behaviour change theories in designing behaviour change interventions, various behaviour change tools, including the Theoretical Domains Framework (TDF), the Behaviour Change Wheel (BCW) and the Behaviour Change Technique Taxonomy (BCTT) have been developed. The TDF is a behavioural framework of 14 domains and considers the individual psychological, social and environmental influences on behaviours (Atkins et al., 2017; Cane et al., 2012; Lawton et al., 2016). The BCW is comprised of three layers to provide a comprehensive and systematic approach to designing behaviour change interventions (Michie et al., 2014). Ninety-three individual BCTs (e.g., instruction on how to perform the behaviour) grouped into 16 categories (e.g., shaping knowledge) comprise the BCTT to identify the ‘observable and replicable’ components of interventions (Michie et al., 2013). Applying behaviour change tools in the design and implementation of complex interventions can assist with understanding an interventions mechanisms of change (Craig et al., 2008; Michie et al., 2014).

Only two studies to-date have applied the TDF to vaccination behaviour. Recently, Kenny et al., (2020) found that the domains of ‘Goals’, ‘Intentions’, ‘Social influences’, and ‘Reinforcement’ could correctly classify whether or not healthcare workers received the influenza vaccine. In addition, Williams et al., (2020) found that the ‘beliefs about consequences’ domain was key in understanding barriers and enablers for receiving a future COVID-19 vaccine in those at high-risk. However, to-date no study has applied the TDF to the understanding of influenza vaccination behaviour amongst the public. Here we address this gap and use the TDF to examine the factors associated with compliance with influenza vaccinations amongst individuals with a chronic respiratory condition. In addition, following the guidance of the UK Medical Research Council on how to develop complex interventions (O’Cathain et al., 2019) we included expert stakeholder engagement, drawing on APEASE criteria, to refine our suggestions for intervention content.

The present study has four key research questions (RQs):

**RQ1**: What are the factors that shape participants’ compliance with influenza vaccination recommendations, and which are the dominant theoretical domains shaping vaccination behaviour?
**RQ2**: What key themes are present within the most dominant theoretical domains and what do these suggest about the tailoring of future intervention content?
**RQ3**: What are the relevant intervention functions and associated behaviour change techniques that we can use to provide potential evidence-based and theoretically informed future intervention content?
**RQ4**: What changes/adaptions do expert stakeholders suggest to ensure intervention content is fit for the future?

## Methods

### Design

This study included qualitative data collected from three sources: open-ended responses from a cross-sectional survey, qualitative data from focus groups, and qualitative data from one to one interviews. All data collection was completed before the start of the COVID-19 pandemic.

### Sample and procedure

Ethics approval for this study was received from the University Ethics Committee. Eligible participants were aged 18-64 years and diagnosed with at least one chronic respiratory condition. Participants were recruited through a variety of methods, including respiratory-focussed charities and associated support groups, print and social media, and a market research company. Our sampling approach sought to recruit participants who exhibited a range of previous vaccination behaviour. As such, we sought to recruit those participants who over the previous five years had always vaccinated, occasionally vaccinated, and those who never vaccinated. We took this approach in order to learn from those people who do regularly comply, those who sometimes comply, and those who do not comply, in order to transfer insights about what does work (enablers) and understand in detail what does not (barriers). The focus group and interview stage of data collection comprised 59 participants, 38 who participated across nine focus groups and 21 in individual interviews. The mean age of participants was 42.8 (SD± 14.8) years and 70% of participants were females. Of these participants, 24% had not vaccinated in the previous five years, 36% had occasionally vaccinated and 41% had always vaccinated, reflecting participants who have a range of compliance with vaccination recommendations. In addition, 101 participants provided responses to the open-ended survey question. Respondents had a mean age of 40.7 (SD: ± 12.8) years and 60% were female. In terms of previous vaccination behaviour, 42% had not received the influenza vaccination in the previous five years, and 47% had occasionally received it.

#### Survey comments

An online cross-sectional survey was used to identify participants’ socio-demographic details, attitudes towards vaccinations (not reported in the current paper), and uptake of the influenza vaccine over the previous five years. Participants could provide a qualitative response to an open-ended question (“*If there are any reasons why you have not received the flu vaccination that the survey has not addressed, please share them below*.”). Participants completed the survey online through Qualtrics.

#### Focus Group and Interviews

Semi-structured focus groups and interviews were conducted using a 13-question interview guide to explore the attitudes of individuals with chronic respiratory conditions towards influenza vaccination. Prompts were also developed for each question to further examine characteristics of influenza vaccine hesitancy. The interview guide was pilot tested with a focus group prior to data collection to ensure question clarity and to determine any changes required to the guide. Participants were sent an email invitation to participate in a focus group and were offered the option of completing an individual interview if they were unable to attend. All focus groups and interviews were facilitated by KD and assisted by AG, and all individual interviews were completed by KD. Audio recordings of focus groups and interviews were transcribed verbatim and anonymised.

#### Stakeholder Meeting

Following the development of initial theory and evidence-informed recommendations for improving influenza vaccination uptake, a meeting was held with six stakeholders to review key recommendations. All stakeholders held key positions within public health and government responsible for the implementation of the influenza vaccination programme in Scotland with remits for policy, strategy and marketing. The APEASE (Affordability, Practicability, Effectiveness/Cost-Effectiveness, Acceptability, Safety and Equity) criteria were applied to each recommendation to help guide discussions to assess the feasibility and appropriateness of the recommendations in addressing the barriers to influenza vaccination uptake in adults with chronic respiratory conditions (Michie et al., 2014).

### Analysis

Figure 1 provides a summary of data analysis procedures used to identify, develop and prioritise recommendations for improving influenza vaccination uptake.

**Figure 1.**
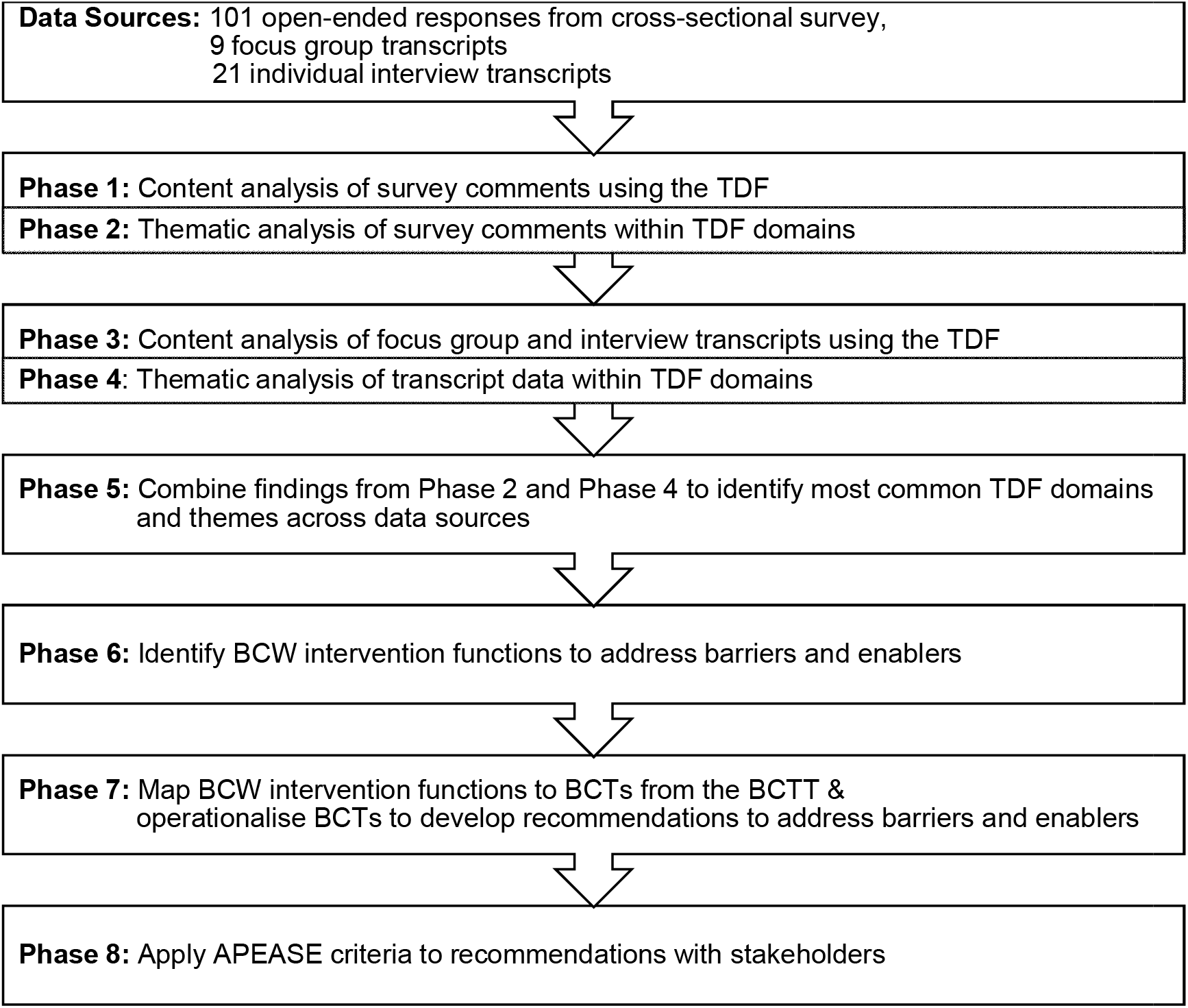
Overview of data analysis procedures and development of recommendations

For RQ1, survey responses were exported from Qualtrics to Microsoft Excel 2016 for analysis. A directed content analysis approach was used (Hsieh & Shannon, 2005), with responses being mapped onto the TDF. Two coders independently reviewed each response and coded it to the most relevant TDF domain. If a response included multiple influences on the uptake of the vaccine, it was reviewed by the coders and mapped to two TDF domains when necessary. The coders met after coding every 20 responses to review coding, identify and discuss any concerns, and to achieve consensus. If coding consensus could not be achieved for a comment, a third team member helped to resolve the discrepancy. Following each meeting, a master copy of the coding was updated to reflect any changes to coding and discrepancy resolutions. Once all responses were coded, 30% of responses were independently reviewed by LW and PF. Consensus in coding was achieved throughout. Thematic analysis was then applied within each TDF domain to determine key themes and subthemes from the survey comments which influenced influenza vaccination behaviours.

Transcripts from the focus groups and interviews were imported into NVivo 12 for analysis (QSR International Pty Ltd., 2020). To conduct a content analysis complementing the one outlined above two coders coded each transcript and mapped participant responses to relevant TDF domains. Quotes could be mapped to two domains where coders deemed it appropriate. Coders met consistently after coding every second focus group transcript and after every third interview transcript to discuss and review coding. Coded files were merged in NVivo and coding stripes were compared. Discrepancies in coding were reviewed and discussed to achieve consensus. When consensus could not be achieved, quotes were passed to LW to resolve. Coding resolutions were saved in the NVivo file. Throughout analysis a codebook was developed to note discussions relating to the TDF and coding associated with each domain. Development of the codebook was an iterative process and was updated following each coding meeting to reflect discussions and coding resolutions. Following coding, LW and PF reviewed 10% of coded quotes to ensure accuracy of coding in relation to the appropriate TDF domain. For RQ2, thematic analysis was applied within each TDF domain to determine the overarching themes and subthemes which influenced influenza vaccination behaviours. Key TDF domains from both the survey comments data and focus group and interview transcripts were identified based on the frequency of content coded under each domain. Major themes and subthemes within domains were ranked based on the number of participants identifying the theme and the number of comments or quotes associated with each theme; this established our sense of which themes were key. Following TDF mapping and thematic analysis of the three streams of qualitative data, data were combined to articulate the commonly identified barriers and enablers from across data these sources.

For RQ3, the key barriers and enablers identified from RQ2 were mapped to the intervention functions level of the BCW. Each factor was compared to the nine intervention functions (e.g., Education, Training, Persuasion) to determine which function(s) would best address the identified barrier or enabler. The BCW intervention functions can be mapped to the BCTT to help identify the ‘active ingredients’ of interventions targeting behaviour change (Michie et al., 2013, 2014). BCTs from the BCTT were identified to address the relevant BCW intervention functions and were operationalised to inform recommendations for intervention content to improve influenza vaccination uptake. This work was conducted by AG and audited by PF and LW.

## Results

### RQ1: What are the factors that shape participants’ compliance with influenza vaccination recommendations, and which are the dominant theoretical domains shaping vaccination behaviour?

Nine of the 14 TDF domains were identified in the open-ended survey responses, with the majority of responses falling under ‘Beliefs about Consequences’ (BACons), followed by the ‘Environmental Context and Resources’ (ECR) and ‘Emotion’ domains. Eight comments were coded to two TDF domains as the responses included multiple influences on influenza vaccination uptake. In the focus groups and interviews, participants’ quotes were mapped to 12 of the 14 TDF domains, with no responses mapped to the ‘Skills’ and ‘Reinforcement’ domains. Barriers and enablers amenable to change were primarily grouped under four domains: ‘BACons’, ‘ECR’, ‘Social Influences’ and ‘Emotion’. As these domains were identified across all data sources they were the focus of subsequent BCW intervention function and BCT mapping.

The full results from behaviour change mapping exercises, operationalised BCT recommendations, and stakeholder meeting feedback can be found in supplementary file 1. The following sections provide a narrative summary of this work.

### RQ2: What key themes are present within the most dominant theoretical domains and what do these suggest about the tailoring of future intervention content?

#### Beliefs about Consequences

Table 1 outlines key findings and supporting quotes from TDF mapping and thematic analysis. Within the BACons domain, participants’ perception of vaccine side effects was the most commonly identified barrier across the data. Following vaccination, participants described experiencing a range of symptoms, from flu-like symptoms and localised pain at the injection site, to significant asthma exacerbations requiring hospitalisation, all of which they attributed to the vaccine. In addition to perceived personal side effects, anecdotal experiences of friends and family who had perceived poor reactions to the vaccine also influenced their decision not to vaccinate.

**Table 1.**
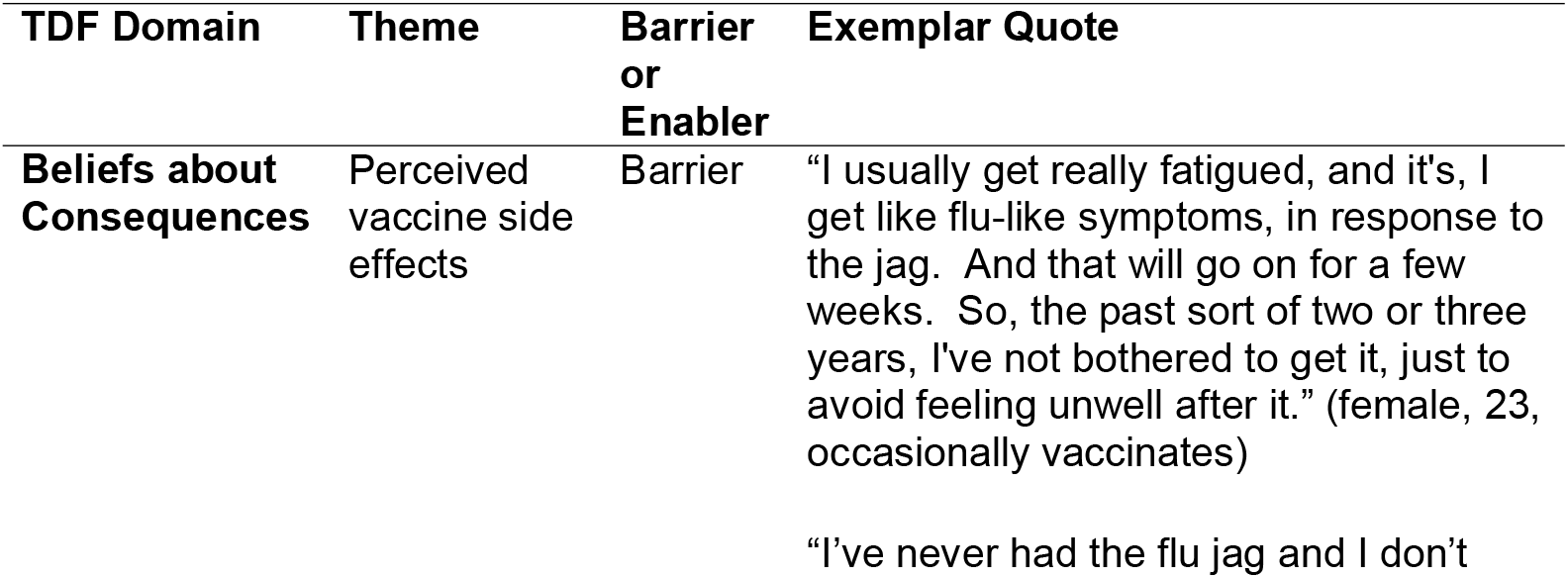
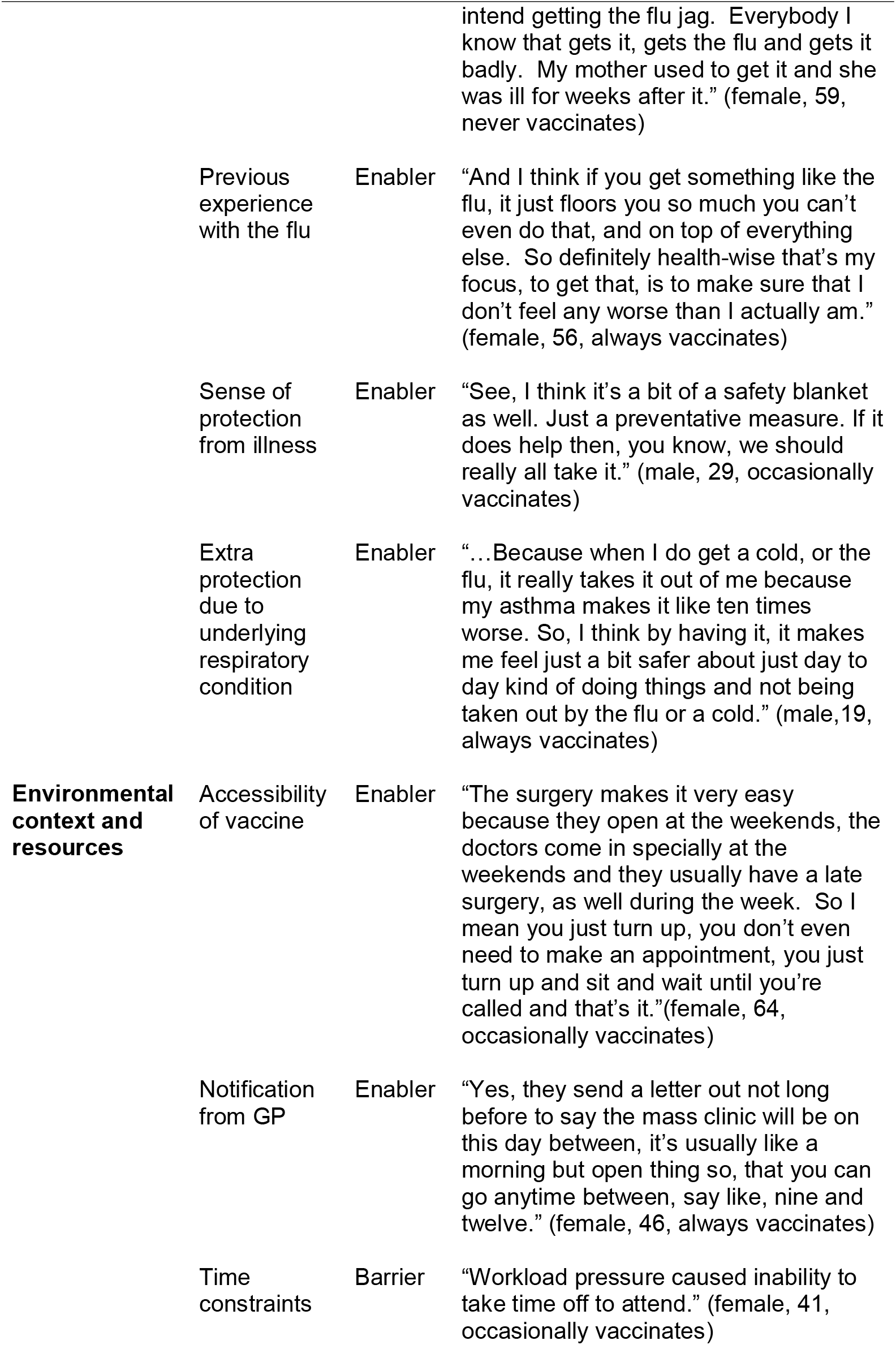
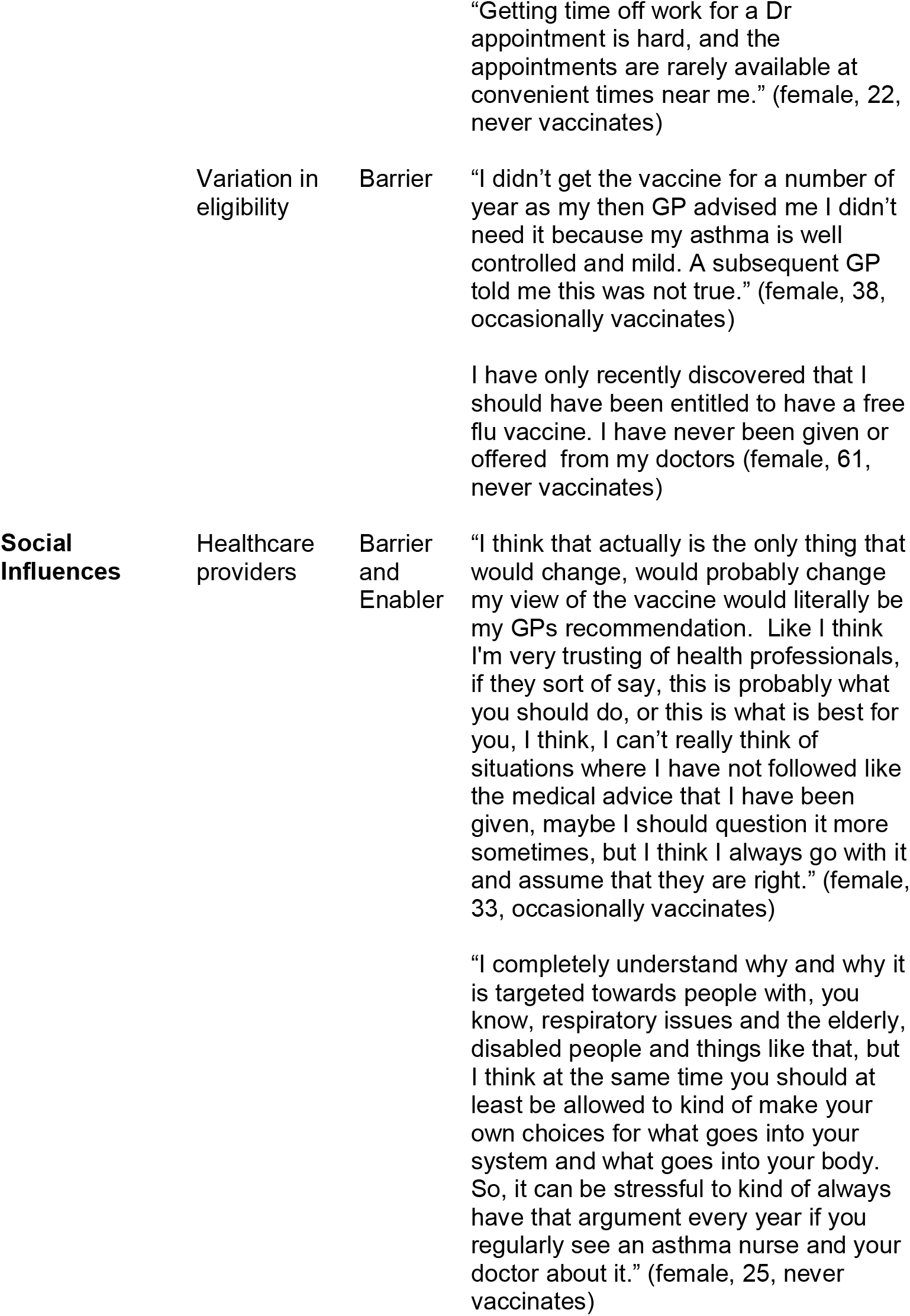
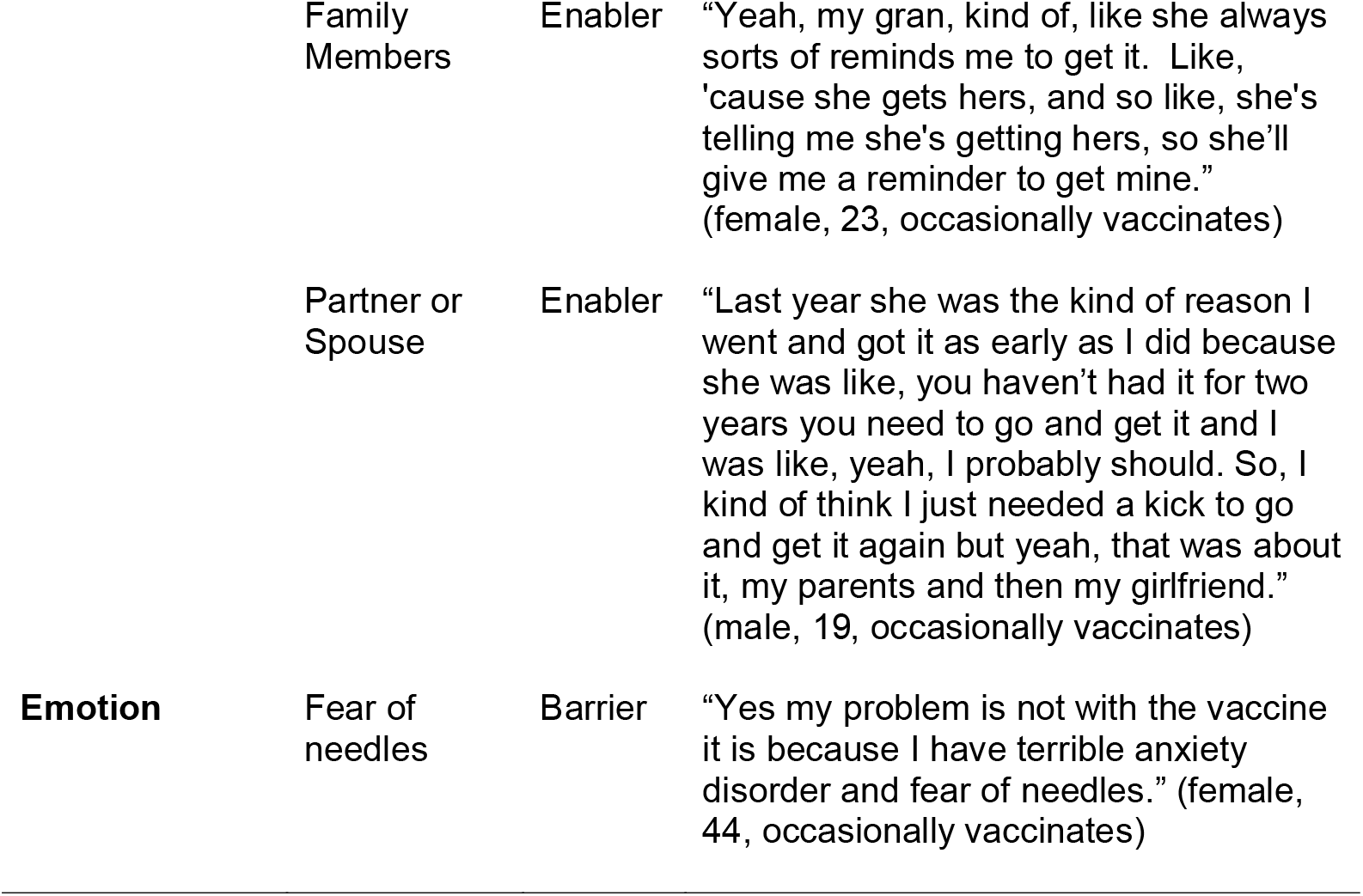
Summary of TDF mapping and thematic analysis

| **TDF Domain** | **Theme** | **Barrier or Enabler** | **Exemplar Quote** |
| --- | --- | --- | --- |
| **Beliefs about Consequences** | Perceived vaccine side effects | Barrier | "I usually get really fatigued, and it's, I get like flu-like symptoms, in response to the jag. And that will go on for a few weeks. So, the past sort of two or three years, I've not bothered to get it, just to avoid feeling unwell after it." (female, 23, occasionally vaccinates)<br>"I've never had the flu jag and I don't<br>intend getting the flu jab. Everybody I know that gets it, gets the flu and gets it badly. My mother used to get it and she was ill for weeks after it." (female, 59, never vaccinates) |
|  | Previous experience with the flu | Enabler | "And I think if you get something like the flu, it just floors you so much you can't even do that, and on top of everything else. So definitely health-wise that's my focus, to get that, is to make sure that I don't feel any worse than I actually am." (female, 56, always vaccinates) |
|  | Sense of protection from illness | Enabler | "See, I think it's a bit of a safety blanket as well. Just a preventative measure. If it does help then, you know, we should really all take it." (male, 29, occasionally vaccinates) |
|  | Extra protection due to underlying respiratory condition | Enabler | "...Because when I do get a cold, or the flu, it really takes it out of me because my asthma makes it like ten times worse. So, I think by having it, it makes me feel just a bit safer about just day to day kind of doing things and not being taken out by the flu or a cold." (male, 19, always vaccinates) |
| **Environmental context and resources** | Accessibility of vaccine | Enabler | "The surgery makes it very easy because they open at the weekends, the doctors come in specially at the weekends and they usually have a late surgery, as well during the week. So I mean you just turn up, you don't even need to make an appointment, you just turn up and sit and wait until you're called and that's it." (female, 64, occasionally vaccinates) |
|  | Notification from GP | Enabler | "Yes, they send a letter out not long before to say the mass clinic will be on this day between, it's usually like a morning but open thing so, that you can go anytime between, say like, nine and twelve." (female, 46, always vaccinates) |
|  | Time constraints | Barrier | "Workload pressure caused inability to take time off to attend." (female, 41, occasionally vaccinates) |
|  |  |  | "Getting time off work for a Dr appointment is hard, and the appointments are rarely available at convenient times near me." (female, 22, never vaccinates) |
|  | Variation in eligibility | Barrier | "I didn't get the vaccine for a number of year as my then GP advised me I didn't need it because my asthma is well controlled and mild. A subsequent GP told me this was not true." (female, 38, occasionally vaccinates)<br>I have only recently discovered that I should have been entitled to have a free flu vaccine. I have never been given or offered from my doctors (female, 61, never vaccinates) |
| **Social Influences** | Healthcare providers | Barrier and Enabler | "I think that actually is the only thing that would change, would probably change my view of the vaccine would literally be my GPs recommendation. Like I think I'm very trusting of health professionals, if they sort of say, this is probably what you should do, or this is what is best for you, I think, I can't really think of situations where I have not followed like the medical advice that I have been given, maybe I should question it more sometimes, but I think I always go with it and assume that they are right." (female, 33, occasionally vaccinates)<br>"I completely understand why and why it is targeted towards people with, you know, respiratory issues and the elderly, disabled people and things like that, but I think at the same time you should at least be allowed to kind of make your own choices for what goes into your system and what goes into your body. So, it can be stressful to kind of always have that argument every year if you regularly see an asthma nurse and your doctor about it." (female, 25, never vaccinates) |
|  | Family Members | Enabler | "Yeah, my gran, kind of, like she always sorts of reminds me to get it. Like, 'cause she gets hers, and so like, she's telling me she's getting hers, so she'll give me a reminder to get mine."<br>(female, 23, occasionally vaccinates) |
|  | Partner or Spouse | Enabler | "Last year she was the kind of reason I went and got it as early as I did because she was like, you haven't had it for two years you need to go and get it and I was like, yeah, I probably should. So, I kind of think I just needed a kick to go and get it again but yeah, that was about it, my parents and then my girlfriend."<br>(male, 19, occasionally vaccinates) |
| **Emotion** | Fear of needles | Barrier | "Yes my problem is not with the vaccine it is because I have terrible anxiety disorder and fear of needles." (female, 44, occasionally vaccinates) |

In contrast to this barrier, three enablers to influenza vaccination uptake were identified within this domain: previous experience of influenza, a sense of protection from illness, and extra protection against the influenza due to underlying health condition. Those who had experienced influenza described their symptoms, including fatigue, body aches and loss of appetite, and that they chose to vaccinate to prevent experiencing this again. Participants also described vaccination provided a sense of protection or ‘safety blanket’ against influenza. This sense of protection was also important due to the high-risk status of participants, who felt influenza further exacerbated issues from their chronic respiratory condition.

#### Environmental Context and Resources

Time and geographical constraints, as well as variation in eligibility for the vaccine were two key barriers within this domain. Participants facing busy schedules and limited GP appointment options made it difficult to prioritise vaccination, especially if they did not work near the practice. Survey participants were more likely to note that they often received confusing advice regarding their eligibility to receive the vaccination for free. Some participants explained they had spent years paying, or being told to pay, for the vaccine as their GP said they were not eligible for the vaccine, only to later learn they had been eligible. This variation in information received from healthcare providers (HCP) limited regular uptake of the vaccine.

Enablers to vaccination included accessibility of the vaccine and receiving notifications encouraged vaccinate within the Environmental Context and Resources domain. Having GP practices offer out of hours appointments, including evenings and weekends, helped ensure people could vaccinate at a convenient time. Participants also found that receiving a notification (e.g., letter, text message or phone call) from their GP practice served as an important reminder to get vaccinated.

#### Social Influences

Three key pillars of social support affected vaccination: healthcare providers (HCP), family members, and partners or spouses. HCP, including GPs and asthma nurses, could help or hinder vaccination. Those who regularly visited or interacted with these providers felt supported and encouraged to vaccinate, while those who did not feel supported were less likely to vaccinate. Support from family members, particularly parents and grandparents, and partners or spouses was also fundamental in encouraging vaccination.

#### Emotion

The main theme identified within the ‘Emotion’ domain was a fear of needles. Comments described a fear of needles and injections, with some explaining the idea of vaccination was enough to trigger an anxiety or panic attack.

### RQ3: What are the relevant intervention functions and associated behaviour change techniques that we can use to provide potential evidence-based and theoretically informed future intervention content?

Across our BCW intervention function analysis of all the TDF domains outlined above, we identified Education (n=8), Persuasion (n=8), Enablement (n=8), Environmental Restructuring (n=4), Modelling (n=2), Training (n=1) and Incentivisation (n=1) as important components of interventions aimed at improving influenza vaccination uptake (Table 2 and Supplementary file 1).

**Table 2.**
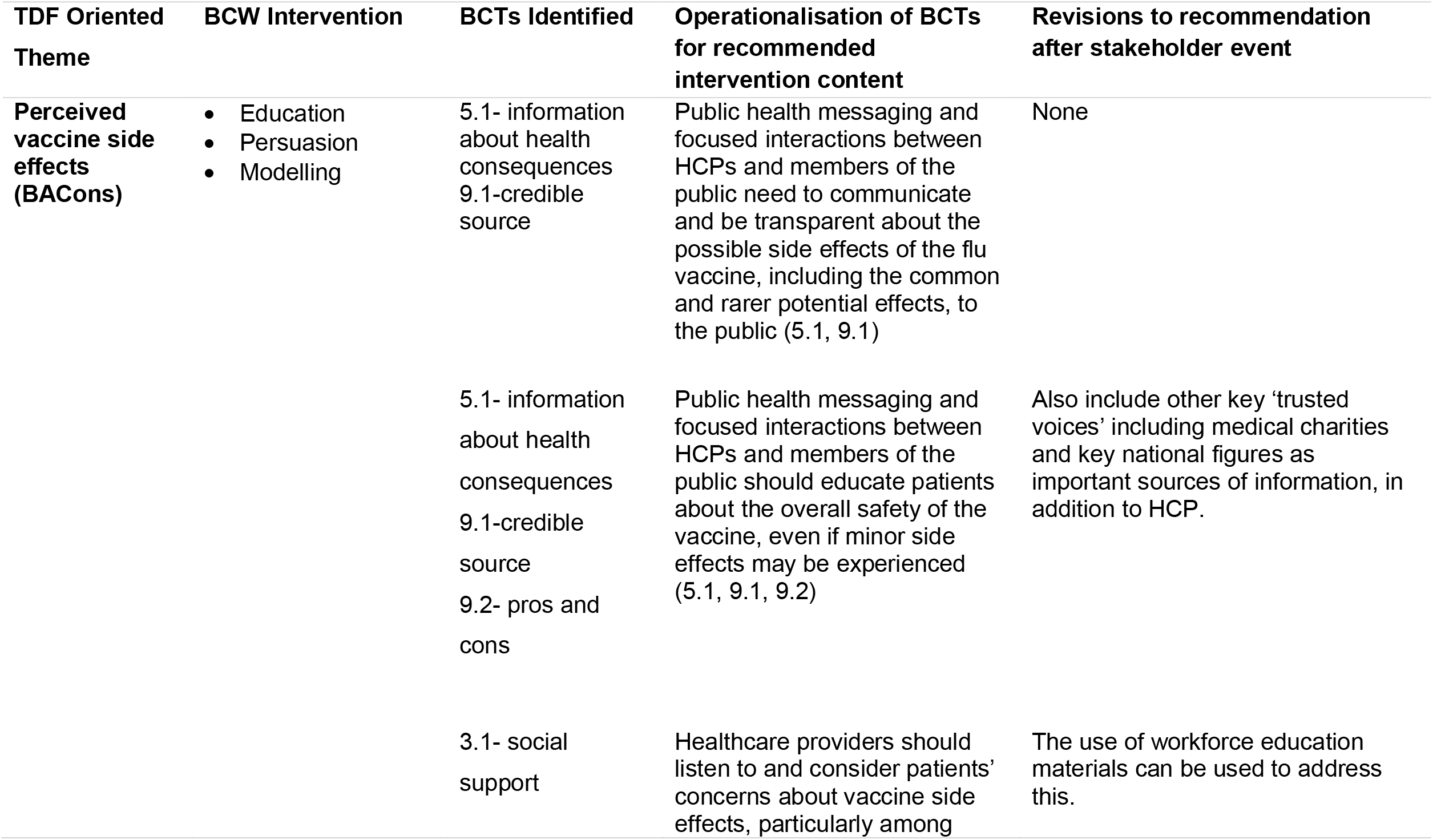
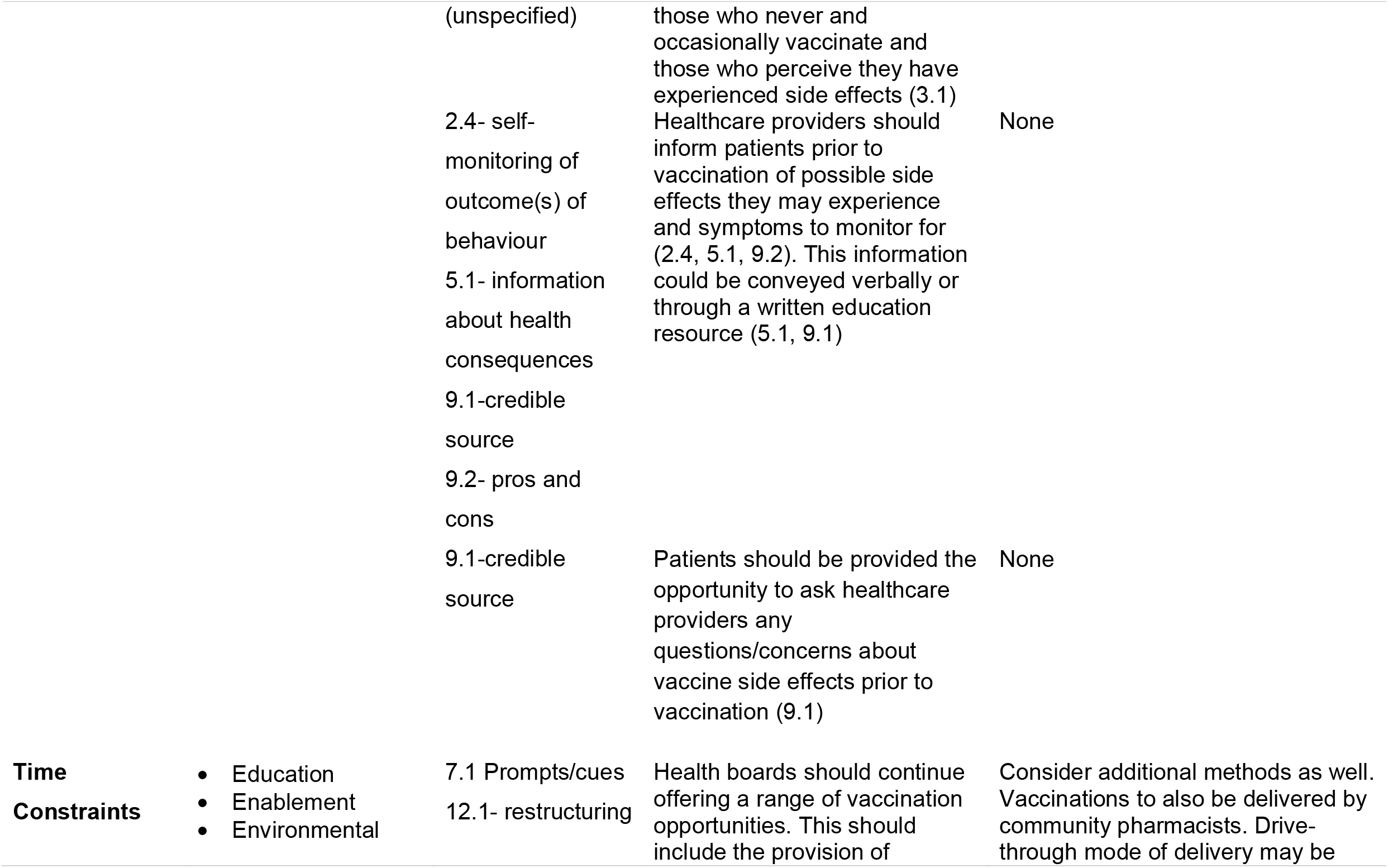
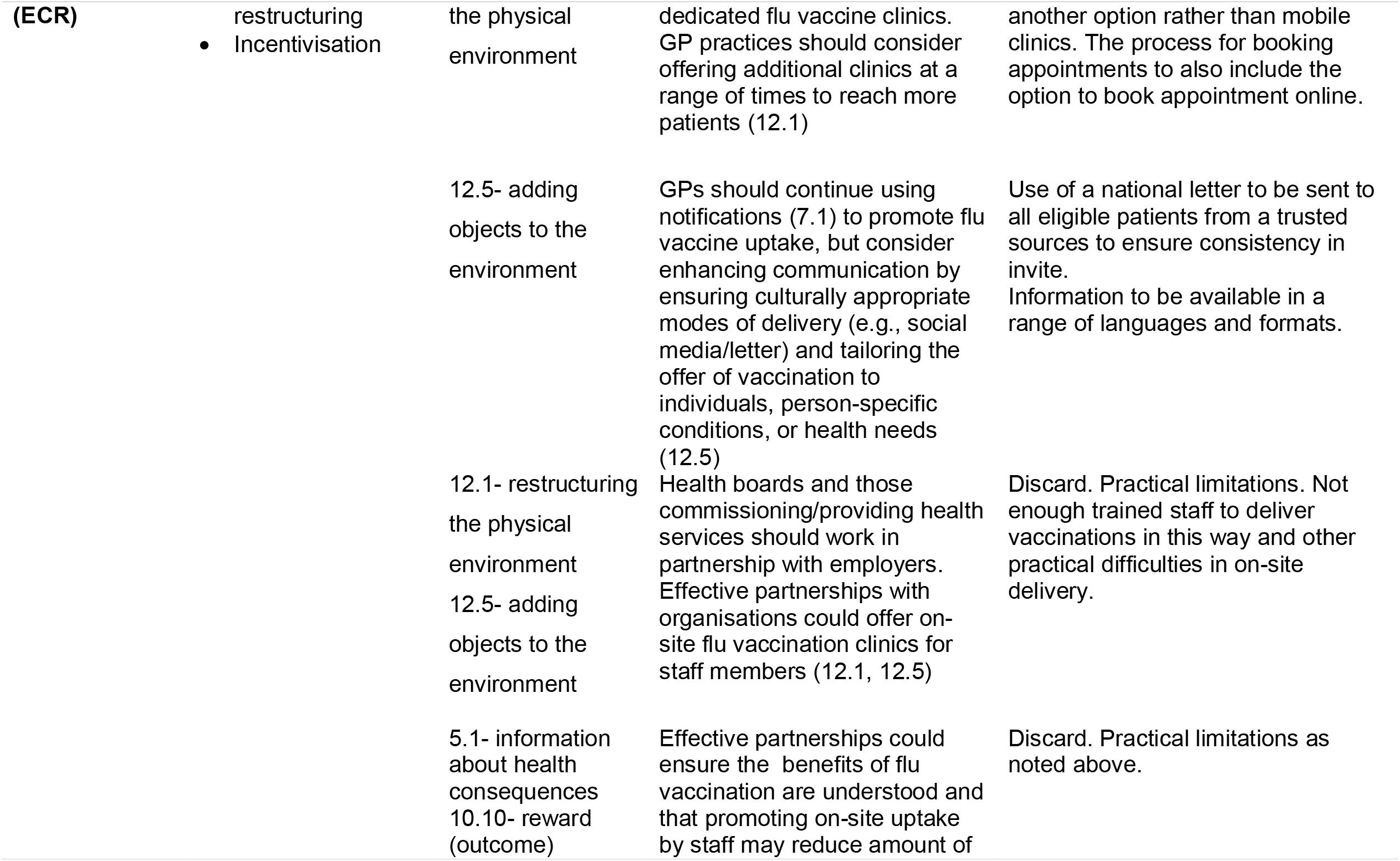
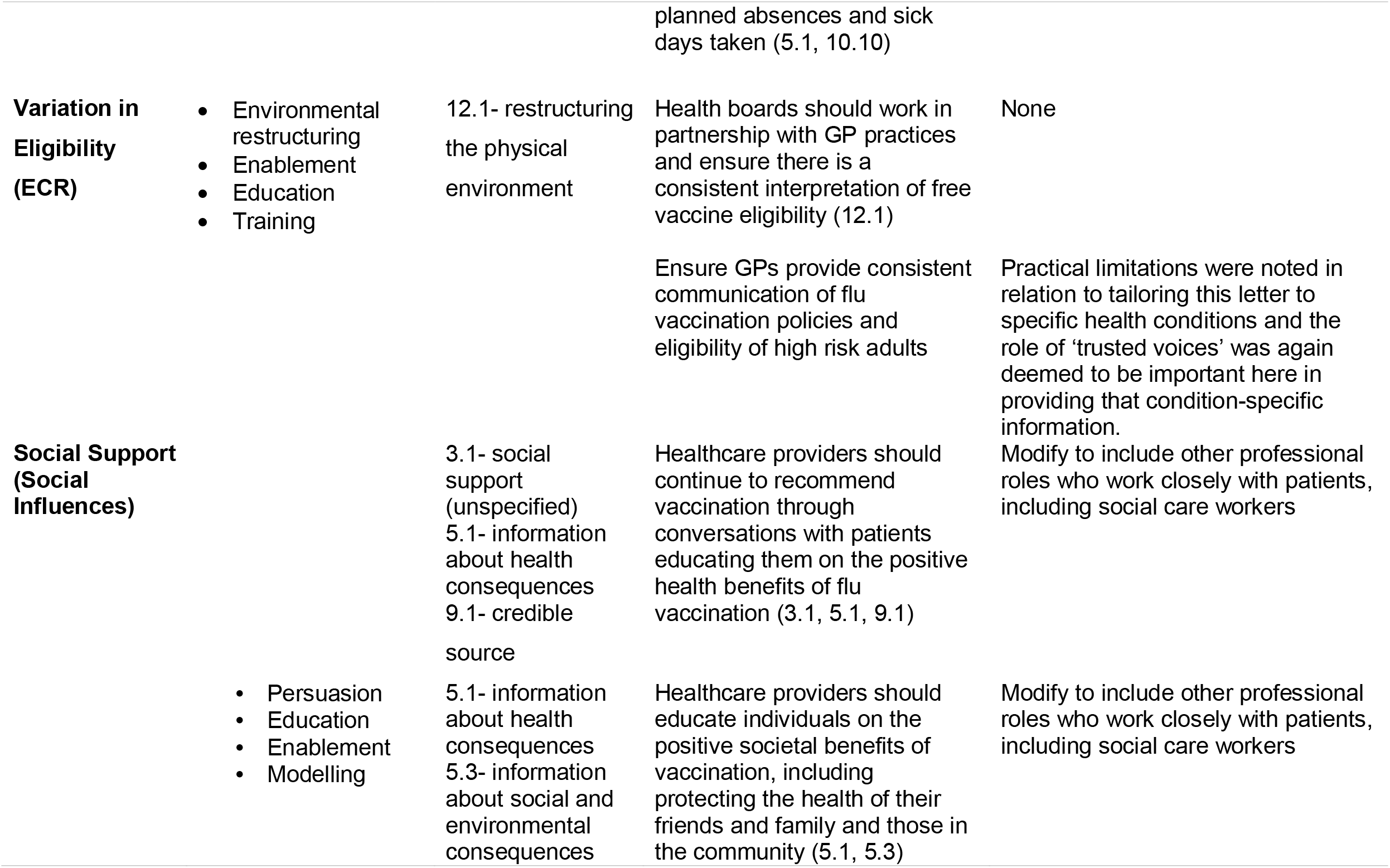
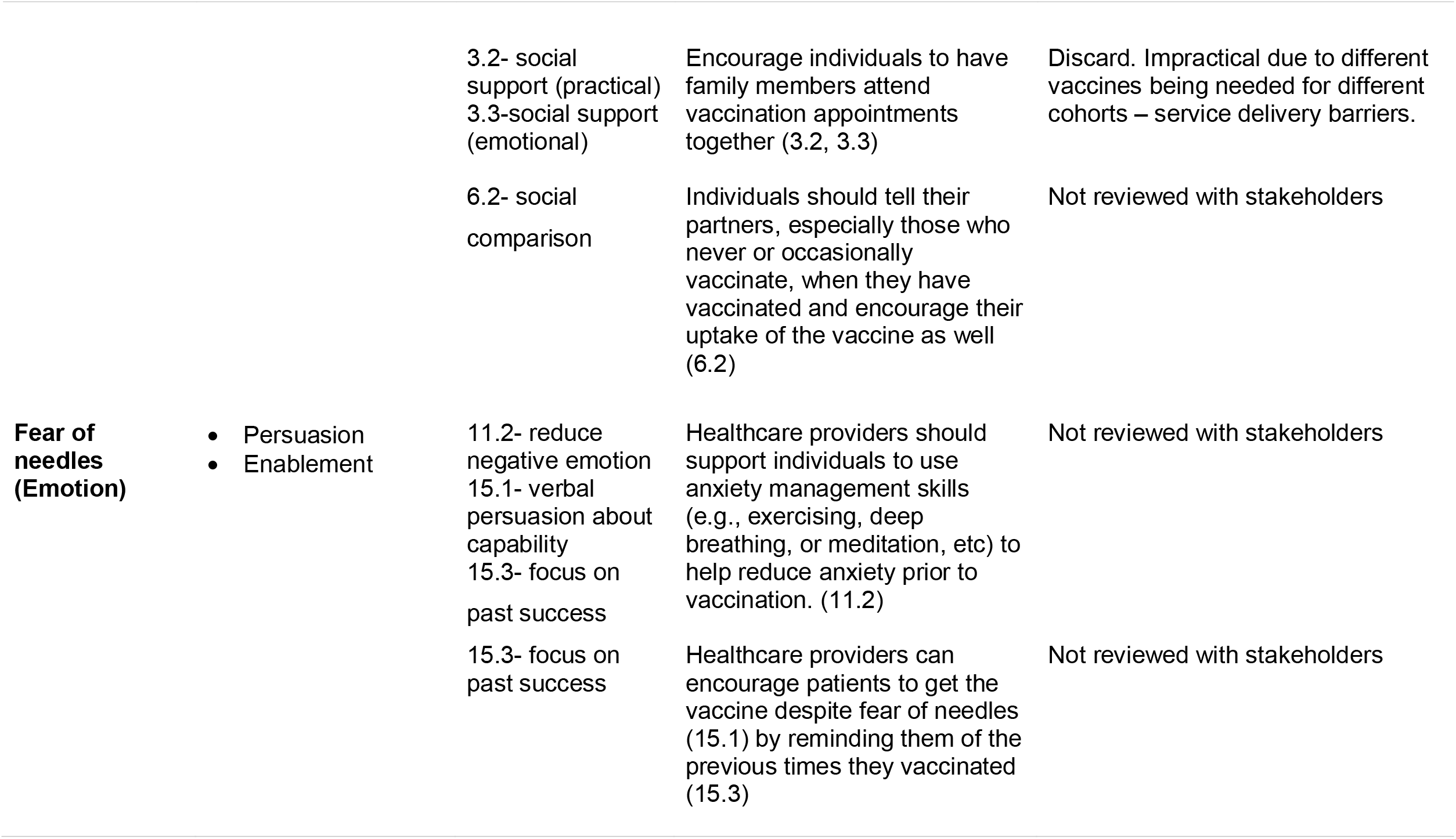
Summary of key TDF oriented themes mapped to intervention functions and individual BCTs

Within the BACons domain, interventions should aim to educate patients about the vaccine and its effects through a range of intervention modalities (e.g., mass media, social media, interactions with HCPs) should be considered to deliver the intervention functions of *Education, Persuasion* and *Modelling*. Overall to directly address BACons, intervention content must redress misinformation, deliver accurate, trustworthy information and provide models of other people’s experiences and thinking in order to educate and persuade. These approaches can be delivered within mass and social media and during HCP and patient interactions. Intervention content must provide frank detail of the reality of side effects including explanations of typical immune responses and how these may feel. Consider training for relevant HCPs to deliver brief behaviour change interventions that educate and persuade those showing vaccine hesitancy within every day interactions. These interactions should feature the same decisional balance outlined above and could draw upon principles of motivational interviewing.

To address the environmental barriers to vaccination, interventions should consider E*nvironmental Restructuring, Enablement, Education and Incentivisation* to encourage vaccine uptake. Overall, interventions should continue to provide vaccination services in GP practices but there needs to be further emphasis on more community-based vaccination clinics. These could include reimbursing those who paid for their vaccination, incentivising workplaces to provide on-site influenza vaccination clinics, enabling more traditional community-based vaccination clinics, or more novel approaches such as a mobile clinics, in addition to out of hours GP vaccination appointments.

Supportive social influences were a positive influence on vaccination behaviour, which *Education, Enablement* and *Persuasion* intervention elements could develop to further encourage uptake. HCP should continue to educate patients on the importance of vaccination and the personal health benefits of vaccinating against influenza, and additional benefits of protecting the health of others by contributing towards herd immunity. Family members and partners could persuade and enable vaccination by attending vaccination clinics together or by using a vaccine ‘buddy system’. Finally, fears of needles and injections could be addressed through persuasion and enablement by supporting the use of anxiety-management techniques before and during vaccination appointments.

Applying the BCTT, we operationalised intervention content recommendations to improve vaccine uptake. Recommendations utilised 22 individual BCTs from 11 of the 16 groupings, with the most commonly identified groups including ‘Natural Consequences’, ‘Social Support’, ‘Antecedents’ and ‘Comparison of Outcomes.’ Table 2 summarises key themes, BCW intervention functions, BCTs applied to our analysis, and how BCTs were operationalised to inform recommendations for intervention content. In Table 2 we present a subset of our recommendations across the key TDF domains. The table in supplementary file 1 presents all of our 26 recommendations in full.

### RQ4: What changes/adaptions do expert stakeholders suruggest to ensure intervention content is fit for the future?

Following the development of recommendations based on the operationalised BCTs, a stakeholder meeting was held with six local stakeholders to review 19 key recommendations against the APEASE criteria (for a full list of the recommendations see supplementary file 1). From this meeting, eight recommendations were considered relevant to help improve vaccine uptake, six were considered to be relevant with slight modifications, while five were considered impractical to implement. Recommendations that were supported included providing public health messaging of vaccine side effects, encouraging conversations between HCP and patients of potential vaccine side effects prior to vaccination, provision of out of hours vaccination appointments at GP practices and other locations, and routinising the information provided to all high-risk patients regarding the vaccine and their eligibility for free vaccination.

Modifications to recommendations included adding other ‘trusted voices’ alongside those of HCP as actors within the recommendations, to provide patients with contextualised information relevant to their condition and potential side effects. These ‘trusted voices’ could include medical charities or national figures, such as the national clinical director, or social care workers. In addition, modifications were suggested to the recommendations relating to vaccine delivery. These included considering other methods such as vaccinations being delivered by community pharmacists, and the potential use of drive-through vaccination centres.

Recommendations deemed impractical by stakeholders due to service-delivery constraints, included providing on-site workplace vaccinations and encouraging a vaccine ‘buddy system’. Worksite vaccination services were considered impractical due the limitations of transporting vaccines and vaccination equipment to workplaces and the availability of trained staff to administer the vaccines. The vaccine ‘buddy system’ recommendation was also considered to be impractical due to the difficulty in having different types of vaccination available to administer at the same time to people from different age cohorts and with different at-risk status.

## Discussion

The present study is the first to apply the BCW and associated BCTT and APEASE tools to the problem of low influenza vaccination uptake among high risk members of the public. Specifically, we triangulate diverse data sources to identify barriers and enablers to seasonal influenza vaccine uptake in adults with chronic respiratory conditions. By applying these tools we were able to conduct a comprehensive analysis of the factors affecting vaccination behaviour, and develop a range of evidence and theory-informed intervention recommendations to improve vaccine uptake. Working with stakeholders direct responsible for the National seasonal influenza vaccination programme a shorter list of actionable recommendations was agreed. Our findings indicate that influenza vaccination is often a complex decision affected by multifaceted individual, social and environmental influences and future intervention components should address this complexity accordingly.

TDF analysis identified ‘Beliefs about Consequences’, ‘Environmental Context and Resources’, ‘Social Influences’ and ‘Emotion’ were the most common TDF domains which affected influenza vaccination. Similarly, Williams et al (2020) also identified ‘Beliefs about Consequences’ to be the most important domain in relation to barriers and enablers to future uptake of a COVID-19 vaccination. However, our findings contrast from those of Kenny et al. (2020) who examined influenza vaccine uptake in HCP. They found that ‘Goals’, ‘Intentions’, ‘Social Influences’ and ‘Reinforcement’ as significant predictors of influenza vaccine uptake in HCPs working in long-term care. Thus suggesting that the barriers to influenza vaccine uptake experienced by HCPs are different to those faced by patients.

Social Influences is the only factor commonly identified across both studies, which demonstrates the importance of social relationships in vaccine uptake for both HCP and patients. This is consistent with research by Gorman et al. (2012) who analysed factors affecting influenza vaccination in pregnant women. They identified women were three times more likely to vaccinate when they were encouraged to by their healthcare provider, while Kenny et al. (2020) found limited peer support decreased influenza vaccination among HCP. Communication between patients and their HCP provides a noteworthy opportunity to support vaccination. HCP should use their time and interactions with patients, and their position as credible sources of health information, to encourage and support vaccination decisions during every day interactions with patients. In addition, supporting a vaccine ‘buddy system’ to encourage friends and families to get vaccinated together may also provide an opportunity to capitalise on the role of social support in vaccination behaviour. During the 2019-2020 influenza season, Public Health England encouraged HCP to use a buddy system in an attempt to reach its goal of 90% vaccination among this group (Ford, 2019). While this recommendation was considered impractical by our stakeholders in Scotland due to the variety of influenza vaccinations available based on age and other risk factors, applying similar systems in communities in other contexts may help address barriers to vaccination and improve uptake.

The findings suggest that perceived vaccine side effects is a predominant barrier to vaccine uptake. Although the influenza vaccine has been shown to be safe and effective, concerns of adverse vaccine events remain (Pebody et al., 2019). Among participants in our study, common side effects including itching and tenderness at the injection site were reported, but infrequent events, including developing influenza, and experiencing eczema and asthma exacerbations, were also attributed to receiving the vaccine. Concerns about vaccine side effects has been well established in influenza vaccine hesitancy research (Schmid et al., 2017; Wiemken et al., 2019), and previous experience with perceived influenza vaccine side effects has also been identified by Ferragut et al. (2020) as a key barrier to vaccination. To help address this concern, HCP should provide open and transparent information about influenza vaccine safety and the prevalence of potential vaccine side effects, from common and less severe symptoms, to the more complex and rare side effects which could be experienced. It is important to provide this information in clear and concise resources, as providing extensive information to those who are vaccine hesitant may have little effect in changing vaccination views or behaviours (Dubé et al., 2020).

Addressing the time and geographical constraints to influenza vaccination is essential in improving uptake of the influenza vaccine. Among our recommendations to offer more out of hour GP appointments during influenza season to improve accessibility of the vaccine, we encourage the use of more traditional community clinics and more novel approaches, such as at workplaces, mobile clinics or drive-through vaccination options. These recommendations to support community vaccination options align with the goals of the Vaccination Transformation Programme in Scotland (VTP; Public Health Scotland, 2020). The VTP was developed by Public Health Scotland in 2017 to support the transition of vaccination services, including influenza vaccinations, away from GP practices and towards community-based options through NHS boards. Our suggestions for transitioning influenza vaccination to community-based routes were largely well received by our stakeholders, although our recommendations for workplace vaccination sites was considered to be impractical as there is limited HCP to conduct this service and difficulty in maintaining vaccine cold chain. The benefits of workplace vaccination have been noted in other countries for improving influenza vaccination rates (Nowalk et al., 2010), and may be useful in some countries for improving accessibility of the vaccine. By providing more accessible vaccination options within communities, it can help address the practical factors to vaccination and allows for more effective use of GP practices and resources.

## Strengths and Limitations

Strengths of this study include the use of multi-modal data collection and subsequent behaviourally informed analysis and expert stakeholder engagement to develop actionable recommendations. We provide a comprehensive picture of the barriers and enablers to uptake of the influenza vaccine in those with a chronic respiratory condition by merging qualitative data from multiple sources. In addition, we also purposively recruited participants from across the vaccine hesitancy continuum meaning that we were able to learn about the barriers and enablers from both regular and non-vaccinating individuals. By reviewing our recommendations against the APEASE criteria with stakeholders, it helped ensure our recommendations are affordable, practical, effective, applicable and equitable feasible to implement into policy and practice. It was a strength of the project that we could recruit a range of professionals directly involved within national provision of vaccines and effectively engage them with the project and its findings. Limitations of the stakeholder engagement stage were that all our stakeholders were recruited from a single national organisation. Further honing of the project’s final recommendations could have been achieved if we had also recruited from the chain of diverse health care professionals involved in vaccination and attempted to gain wider perspectives from other trusted sources such as the third sector. Other limitations include that our findings are limited to participants living in Scotland with a chronic respiratory condition, and so may not be generalizable to other chronic health conditions or national contexts. Furthermore data were collected before the onset of COVID-19 and it may be that beliefs and behaviours about vaccination have changed radically as a result of this pandemic.

## Conclusion

This study triangulates diverse findings to conduct a comprehensive behavioural diagnosis of compliance with influenza vaccination advice. With the help of stakeholder engagement we have shown that the factors affecting vaccine uptake were multifaceted and multileveled. They could be theorised as relating to the TDF domains of ‘Beliefs about Consequences’, ‘Environmental Context and Resources’, ‘Social Influences’ and ‘Emotion’. The study suggested a suite of complementary multi-level intervention components may be most useful to enhance vaccination compliance involving a range of diverse actors, intervention recipients and settings. Mass and social media interventions, and interactions between recipients and HCP should include clear and concise information about vaccine side-effects and directly address misinformation. Community-based vaccination delivery methods should be enhanced by modifying traditional and adopting novel approaches.

## Data Availability

The data are not publicly available due to privacy or ethical restrictions.

## Data availability statement

The data are not publicly available due to privacy or ethical restrictions.

## Acknowledgements

This study was funded by the Chief Scientist Office in Scotland (reference: HIPS 18/37). The authors would like to thank Dana O’Flynn for assisting with preliminary TDF coding of survey comments. We would also like to thank the stakeholders who participated in our stakeholder meeting for sharing their valuable insights and recommendations.

