## Supplementary File 1 for "Barriers and enablers to influenza vaccination uptake in adults with chronic respiratory conditions: Applying the behaviour change wheel to specify multi-levelled tailored intervention content"

| **TDF Domain** | **Domain subthemes** | **Focus group data** | | **# survey comments (n=101)** | **Exemplar quotes** (gender/age/vaccine uptake in the past five years) | **BCW intervention functions** | **Identified BCTs** | **Operationalised BCT for recommended intervention content** | **Stakeholder meeting** | |
| --- | --- | --- | --- | --- | --- | --- | --- | --- | --- | --- |
|  |  | **# Participants (n=59)** | **# Quotes** |  |  |  |  |  | Stakeholder Feedback (keep, modify, discard) | Notes and Revisions to recommendation |
| **Beliefs about Consequences** | Perceived vaccine side effects (experienced by self or others) | 26 | 56 | 21 | I usually get really fatigued, and it's, I get like flu-like symptoms, in response to the jag. And that will go on for a few weeks. So, the past sort of two or three years, I've not bothered to get it, just to avoid feeling unwell after it. (female, 23, occasionally vaccinates)  I’ve never had the flu jag and I don’t intend getting the flu jag. Everybody I know that gets it, gets the flu and gets it badly. My mother used to get it and she was ill for weeks after it. (female, 59, never vaccinates)  No, it was solely in relation to the eczema and there was a definite correlation, like every year you get towards flu jag season in November-ish, October, November. I’d go to the doctor, get the jag and then three to four weeks later I’d have a massive flare-up of my eczema. And it was several years in a row it was exactly the same. (male, 39, never vaccinates) | - Education - Persuasion - Modelling | - 2.4- Self-monitoring of outcome(s) of behaviour - 3.1- social support (unspecified) - 5.1- information about health consequences - 5.2- salience of consequences - 9.1-Credible source - 9.3- pros and cons | 1. Public health messaging and focused interactions between HCPs and members of the public need to communicate and be transparent about the possible side effects of the flu vaccine, including the common and rarer potential effects, to the public (5.1, 9.1) | Keep |  |
|  |  |  |  |  |  |  |  | 2. Public health messaging and focused interactions between HCPs and members of the public should educate patients about the overall safety of the vaccine, even if minor side effects may be experiences (5.1, 9.1, 9.2) | Modify | Also include other key ‘trusted voices’ including medical charities and key national figures as important sources of information, in addition to HCP. |
|  |  |  |  |  |  |  |  | 3. Healthcare providers should listen to and consider patients’ concerns about vaccine side effects, particularly among those who never and occasionally vaccinate and those who perceive they have experienced side effects (3.1). | Keep | The use of workforce education materials can be used to address this. |
|  |  |  |  |  |  |  |  | 4. Healthcare providers should inform patients prior to vaccination of possible side effects they may experience and symptoms to monitor for (2.4, 5.1, 9.2). This information could be conveyed verbally or through a written education resource (5.1, 9.1) | Keep |  |
|  |  |  |  |  |  |  |  | 5. Patients should be provided the opportunity to ask healthcare providers any questions/concerns about vaccine side effects prior to vaccination (9.1) | Keep |  |
|  | Personal experience with the flu | 26 | 49 | 5 | It was just really dreadful. And I didn’t probably get over it for about a month or so, a good three weeks. Even going back to work, I was kind of knackered by the end of the day. It’s just like full body exhaustion and, you know, aches and pains for weeks after. (female, 25, never vaccinates)  And I think if you get something like the flu, it just floors you so much you can’t even do that, and on top of everything else. So definitely health-wise that’s my focus, to get that, is to make sure that I don’t feel any worse than I actually am. (female, 56, always vaccinates) | - Education - Persuasion | 5.1-information about health consequences  5.5- anticipated regret  9.2- pros and cons OR 9.3- comparative imagining of future outcomes? | 6. Public health messaging and focused interactions between HCPs and members of the public should support vaccine uptake by providing education about flu vaccine effectiveness in preventing the flu and lessening flu symptoms (5.1) | Not reviewed with stakeholders |  |
|  | Protection from illness | 31 | 57 | - | See, I think it’s a bit of a safety blanket as well. Just a preventative measure. If it does help then, you know, we should really all take it. (male, 29, occasionally vaccinates)  I'm a bit…yeah, sorry, like, I had it for the first time recently, and it was the best. I didn’t get ill for months. Which usually I get ill, like, every month, so obviously, it affects my asthma. But yeah, it was great. (female, 21, occasionally vaccinates)  I think it’s amazing… I grew up having colds, flus and that all my life so when I got to the stage with my health problems I started getting the flu jag it’s been sort of amazing. I’ve not had a really bad flu. (female, 50, always vaccinates) | - Education - Persuasion | 5.1-information about health consequences | 7. Public health messaging and focused interactions between HCPs and members of the public should support vaccine uptake by providing education about flu vaccine effectiveness in preventing the flu and lessening flu symptoms (5.1) | Not reviewed with stakeholders |  |
|  | Extra protection with underlying respiratory condition | 20 | 30 | 2 | I don’t know, it just kind of in my head it means that I won’t, I possibly won’t get the flu. Because when I do get a cold, or the flu, it really takes it out of me because my asthma makes it like ten times worse. So, I think by having it, it makes me feel just a bit safer about just day to day kind of doing things and not being taken out by the flu or a cold. Stuff like that. (male, 19, always vaccinates)  I've got asthma, and I find that normally every winter I will have a chest infection, and without the flu jab it would be so much worse, it floors me as it is, but I would be so much more ill if I didn't have that flu jab. (female, 43, always vaccinates) | - Education - Persuasion | 5.1-information about health consequences  5.5- anticipated regret  5.2- salience of consequences  9.3- comparative imagining of future outcomes | 8. Public health messaging and focused interactions between HCPs and members of the public should support vaccine uptake by providing education about flu vaccine effectiveness in preventing the flu and lessening flu symptoms (5.1). Consider showing the severity of effects amongst those most vulnerable (5.5, 5.2, 9.3) | Not reviewed with stakeholders |  |
| Environmental Context and Resources | Accessibility of vaccine | 33 | 57 | 4 | the surgery makes it very easy because they open at the weekends, the doctors come in specially at the weekends and they usually have a late surgery, as well during the week. So I mean you just turn up, you don’t even need to make an appointment, you just turn up and sit and wait until you’re called and that’s it. (female, 64, occasionally vaccinates)  They’ve certainly got, I think, days when they do it, they have a flu clinic or whatever they would call them, and try and put you into that. But if you say, no, I can’t really make that, you just make an appointment with the practice nurse, so there’s flexibility. (female, 55, always vaccinates) | - Enablement | - 12.1- restructuring physical environment | 9. Health boards should continue offering a range of opportunities to increase vaccination. This should include the provision of dedicated community and mobile flu vaccine clinics. GP practices should consider offering additional clinics at a range of times to reach more patients (12.1) | Modify | Consider additional methods as well. Vaccinations to also be delivered by community pharmacists. Drive-through mode of delivery may be another option rather than mobile clinics. The process for booking appointments to also include the option to book appointment online. |
|  | Notification from GP | 22 | 31 | 2 | Yes, they send a letter out not long before to say the mass clinic will be on this day between, it’s usually like a morning but open thing so, that you can go anytime between, say like, nine and twelve (female, 46, always vaccinates) | - Environmental restructuring - Enablement | - 7.1- Prompts/cues - 12.5- adding objects to the environment | 10. GPs should continue using notifications (7.1) to promote flu vaccine uptake, but consider enhancing communication by ensuring culturally appropriate modes of delivery (e.g., social media/letter) and tailoring the offer of vaccination to individuals, person-specific conditions, or health needs (12.5) | Modify | Use of a national letter to be sent to all eligible patients from a trusted sources to ensure consistency in invite.  Information to be available in a range of languages and formats. |
|  | Inaccessible/Time constraints | 18 | 28 | 10 | Workload pressure caused inability to take time off to attend. (female, 41, occasionally vaccinates)  Getting time off work for a Dr appointment is hard, and the appointments are rarely available at convenient times near me. (female, 22, never vaccinates) | - Education - Enablement - Environmental restructuring - Incentivisation | 5.1- Information about health consequences  10.10- reward (outcome)  12.1- restructuring the physical environment  12.5- adding objects to the environment | 11. All GP practices should continue offering a range of opportunities to increase vaccination. This should include the provision of dedicated flu vaccine clinics. GP practices should consider offering additional clinics at a range of times to reach more patients (12.1) | Keep |  |
|  |  |  |  |  |  |  |  | 12. Health boards and those commissioning/providing health services should work in partnership with employers. Effective partnerships with organisations could offer on-site flu vaccination clinics for staff members (12.1, 12.5) | Discard | Practical limitations. Not enough trained staff to deliver vaccinations in this way and difficulties in bringing vaccination equipment on-site. |
|  |  |  |  |  |  |  |  | 13. Effective partnerships could ensure the benefits of flu vaccination are understood and that promoting on-site uptake by staff may reduce amount of planned absences and sick days taken (5.1, 10.10) | Discard | Practical limitations. Not enough trained staff to deliver vaccinations in this way and difficulties in bringing vaccination equipment on-site. |
|  |  |  |  |  |  |  |  | 14. Health boards should consider offering vouchers/ reimbursements for vaccination costs to eligible individuals who pay for the vaccination at pharmacies (12.5) | Keep |  |
|  | Variation in eligibility | 3 | 3 | 17 | During some years I have received confusing advice about whether I'm eligible to have it free of charge (female, 39, always vaccinates)  I have only recently discovered that I should have been entitled to have a free flu vaccine. I have never been given or offered from my doctors (female, 61, never vaccinates)  I didn’t get the vaccine for a number of year as my then GP advised me I didn’t need it because my asthma is well controlled and mild. A subsequent GP told me this was not true. (female, 38, occasionally vaccinates) | - Environmental restructuring - Enablement - Education - Training | 12.1- restructuring the physical environment  5.1-Information about health consequences  8.1- behaviour practice/rehearsal  8.3- habit formation | 15. Health boards should work in partnership with GP practices and ensure there is a consistent interpretation of free vaccine eligibility (12.1) | Keep |  |
|  |  |  |  |  |  |  |  | 16. Ensure GPs provide consistent communication of flu vaccination policies and eligibility of high risk adults (12.1) | Modify | Practical limitations were noted in relation to tailoring this letter to specific health conditions and the role of ‘trusted voices’ was again deemed to be important here in providing that condition-specific information. |
|  |  |  |  |  |  |  |  | 17. GPs and healthcare providers should routinise the informing of all high risk patients every year (8.1 GP, 8.3 GP) | Keep |  |
| Social Influences | Health Care Professionals | 22 | 40 | 1 | you go and you do it, the nurse is always really nice and jolly and you usually get a good laugh when you go, bit of banter with the nurse, and they always make you feel so comfortable. (female, 56, always vaccinates) | - Persuasion - Education | - 3.1- social support (unspecified) - 5.1- Information about health consequences - 5.3- information about social and environmental consequences - 9.1- Credible source | 18. Healthcare providers should continue to recommend vaccination through conversations with patients educating them on the positive health benefits of flu vaccination (3.1, 5.1, 9.1) | Modify | Modify to include other professional roles who work closely with patients, including social care workers |
|  |  |  |  |  |  |  |  | 19. Healthcare providers should educate individuals on the positive societal benefits of vaccination, including protecting the health of their friends and family and those in the community (5.1, 5.3) | Modify | Modify to also include other professional roles who work closely with patients, including social care workers |
|  | Family members | 11 | 18 | 1 | Yeah, my gran, kind of, like she always sorts of reminds me to get it. Like, 'cause she gets hers, and so like, she's telling me she's getting hers, so she’ll give me a reminder to get mine. (female, 23, occasionally vaccinates) | - Enablement - Persuasion | - 3.2- social support (practical) - 3.3-social support (emotional) - 6.2- Social comparison - 6.3- Information about others’ approval | 20. Encourage individuals to have family members attend vaccination appointments together (3.2, 3.3) | Discard | Impractical due to different vaccines being needed for different cohorts – service delivery barriers. |
|  |  |  |  |  |  |  |  | 21. Family members may want to encourage vaccine uptake by others by noting when they and other family members have vaccinated (6.2, 6.3) | Not reviewed with stakeholder |  |
|  |  |  |  |  |  |  |  | 22. Healthcare providers should promote family vaccinations, so all family members can get vaccinated at one appointment. | Discard /modify | Impractical, but there may be a role for opportunistic vaccination with vaccination groups (e.g., pregnant mum taking child to vaccine appointment) |
|  | Partner/Spouse | 7 | 10 | 1 | Again, my parents when I was younger and then my girlfriend who has worse asthma than I do. Last year she was the kind of reason I went and got it as early as I did because she was like, you haven’t had it for two years you need to go and get it and I was like, yeah, I probably should. (male, 19, occasionally vaccinates) | - Enablement - Modelling | - 3.2- social support (practical) - 3.3-social support (emotional) - 6.2- Social comparison | 23. Encourage individuals to book and attend vaccination appointments with partner/spouse (3.2, 3.3) | Discard | Impractical due to different vaccines being needed for different cohorts – service delivery barriers. |
|  |  |  |  |  |  |  |  | 24. Individuals should tell their partners, especially those who never or occasionally vaccinate, when they have vaccinated and encourage their uptake of the vaccine as well (6.2). | Not reviewed with stakeholders |  |
| Emotion | Fear of needles | 11 | 16 | 8 | Yes my problem is not with the vaccine it is because I have terrible anxiety disorder and fear of needles. (female, 44, occasionally vaccinates) | - Persuasion - Enablement | - 11.2- reduce negative emotions - 15.1- verbal persuasion about capability - 15.3- focus on past success | 25. Healthcare providers should support individuals to use anxiety management skills (e.g., exercising, deep breathing, or meditation, etc) to help reduce anxiety prior to vaccination. (11.2) | Not reviewed with stakeholders |  |
|  |  |  |  |  |  |  |  | 26. Healthcare providers can encourage patients to get the vaccine despite fear of needles (15.1) by reminding them of the previous times they vaccinated (15.3) | Not reviewed with stakeholders |  |
